# Cost-effectiveness of measles rapid diagnostic tests for replacing or expanding laboratory testing in Ethiopia

**DOI:** 10.64898/2026.06.15.26355722

**Authors:** Fenella McAndrew, Kaungmyat Khant, Dominic Delport, Romesh G Abeysuriya, Mikiyas Alayu, Freya JI Fowkes, Riccardo Gavioli, Kiddus Yitbarek Kehali, Mirchaye Mekoro, Kerryn A Moore, Meru Sheel, Basleal Yirgu, Win Han Oo, Nick Scott

**Affiliations:** Burnet Institute, Melbourne, Australia; Department of Epidemiology and Preventive Medicine, Monash University, Melbourne, Victoria, Australia; Centre for Epidemiology and Biostatistics, Melbourne School of Population & Global Health, University of Melbourne, Victoria, Australia; Sydney Infectious Diseases Institute, Faculty of Medicine and Health, The University of Sydney; Sydney School of Public Health, Faculty of Medicine and Health, The University of Sydney; School of Public Health, Faculty of Health, University of Technology Sydney, Ultimo, New South Wales, Australia; Department of Health Policy and Management, Faculty of Public Health, Jimma University, Jimma, Ethiopia; Health Poverty Action, Adidas Ababa, Ethiopia; Ethiopian Public Health Institute, Addis Ababa, Ethiopia

**Keywords:** agent-based model, cost-effectiveness, measles, outbreak, rapid diagnostic tests

## Abstract

**Background:** In low- and middle-income countries, laboratory testing to rapidly detect measles outbreaks is limited by infrastructure availability and high costs. This study estimates the potential impact and cost-effectiveness of measles rapid diagnostic tests (RDTs) if implemented nationally in Ethiopia to either replace or expand current testing.

**Methods:** An agent-based model to simulate measles outbreaks was calibrated to Ethiopian measles surveillance data. Modelled outbreak outcomes were aggregated over a 10-year period. Scenarios included using RDTs to (1) replace laboratory testing; (2) replace epidemiological linkage; and (3) increase case detection, in addition to replacing laboratory testing and epidemiological linkage. Testing and outbreak response costs (in 2025 US$) were obtained from Ethiopian Public Health Institute from a government perspective. Total costs and disability-adjusted life years (DALYs) for each scenario were compared to baseline.

**Results:** All scenarios were cost saving compared to baseline. Replacing laboratory testing with RDTs saved US$4.2M (3.2M-4.9M) over 10-years, but due to very low testing rates the benefits of eliminating laboratory testing delays were offset by missed cases from the lower RDT sensitivity, leading to similar outbreak detection times and DALYs. Replacing epidemiological linkage with RDTs had similar DALYs but increased the cost savings to US$9.7M. Using RDTs to double case detection reduced outbreak detection time from 113 to 80 days, averted 17,000 DALYs, and saved US$4.3M.

**Conclusions:** In Ethiopia, use of measles RDTs could be cost saving, and if used to expand testing could prevent measles infections through faster outbreak detection and response.

**Key messages:** *What is already known on this topic:* Laboratory testing is the main surveillance tool for detection of measles outbreaks, but in low- and middle-income countries it is relatively expensive and there can be long delays in getting results due to logistic and capacity constraints in some locations.

*What this study adds:* Rapid diagnostic tests (RDTs) for measles have been developed to lower testing costs and reduce measles outbreak response times. This analysis used an agent-based model to simulate measles outbreaks in Ethiopian woredas and assess the cost-effectiveness of introducing RDTs for enhanced measles surveillance.

*How this study might affect research, practice or policy:* The findings of this study provide evidence that deploying measles RDTs in low- and middle-income countries such as Ethiopia where laboratory testing capacity is limited could be cost-saving and reduce disease burden.

## 1 Background

Measles is a highly infectious disease that spreads through the air and can result in severe disease or death and is most common in children [1]. It has been identified as a high priority disease by the World Health Organization (WHO) and Member States in all WHO Regions have adopted measles elimination goals [2]. Despite the commitments of WHO Member States to elimination and the availability of a safe and effective vaccine, measles continues to claim lives among young children worldwide due to gaps in vaccination coverage and limitations of systems to detect and respond to outbreaks [3].

A strong surveillance system is required to rapidly detect and respond to measles outbreaks. Currently laboratory testing using Enzyme-Linked Immunosorbent Assay (ELISA) is the main surveillance tool for detection of measles outbreaks [4], but in low- and middle-income countries it is relatively expensive, testing coverage is limited by the availability of infrastructure, and there can be long delays in getting results due to logistic and capacity constraints in some locations. Rapid diagnostic tests (RDTs) for measles have the potential to overcome some of these challenges. As of 2024 there were no RDTs commercially available for measles [5], but clinical and now pilot implementation studies have occurred or are underway, for example in Zimbabwe, Malaysia, Brazil, and Ethiopia [6-8]. Use of measles RDTs for surveillance could potentially be cheaper than laboratory testing and could minimise delays in outbreak detection and enable expanded testing coverage. However, measles RDTs may have lower sensitivity and specificity than laboratory testing using ELISA (RDT: sensitivity 0.9, specificity 0.95; ELISA: sensitivity 0.907, specificity 0.96) [8, 9]. When considering the introduction of measles RDTs as either a replacement or expansion of laboratory testing, the trade-offs between lower cost, higher coverage and lower sensitivity and specificity can be considered in a health economic analysis.

In 2024 Ethiopia reported 31,044 measles cases, the third highest of any country in that year behind only Democratic Republic of the Congo and Iraq [10]. However, this is likely to be an underestimate of the true number of infections due to limited coverage of laboratory testing [11]. At a national level measles in Ethiopia is endemic and presents as ongoing case detection. However, Ethiopia is made up of approximately 1050 administrative districts known as woredas, and at a woreda level measles presents as collections of outbreaks which are responded to by local public health units. An outbreak in a woreda is declared if there are three laboratory-confirmed cases within a 30-day period [12]. Once an outbreak is declared, further cases are detected through epidemiological linkage in preference of laboratory testing. This approach relies on dedicated teams to identify contacts of confirmed measles cases who meet the clinical case definition, which increases case detection speed but may have low sensitivity and specificity and can result in uncertain knowledge of when the outbreak has ended [13]. The outbreak response often includes supplementary vaccinations given to children aged six months to ten years, although stockouts and other logistical constraints means that this is not always possible.

This study aims to use modelling to estimate the potential impact and cost-effectiveness of RDTs if they were implemented nationally in Ethiopia to either: replace laboratory testing (eliminating delays associated with processing results); replace both laboratory testing and epidemiological linkage (increasing the sensitivity and specificity of case detection following outbreak detection and facilitating faster outbreak responses); or increase testing coverage (hence reducing outbreak detection and response time) as well as replacing laboratory testing and epidemiological linkage. These outcomes are relevant to other countries with a high measles burden and limited laboratory testing capacity.

## 2 Methodology

### 2.1 Setting

Ethiopia had an estimated population of 135 million in 2025 [14] and is geographically subdivided into regions (level 1), zones (level 2) and woredas (level 3). There are approximately 1050 woredas, which each have between 5,337 and 609,401 people (median = 87,230). This study considers measles outbreaks defined at a woreda level to align with public health responses.

### 2.2 Data

The Ethiopian Public Health Institute surveillance system includes line-listed data for all probable cases of measles or rubella nationally. For the study purpose, the national surveillance dataset between 2011 and 2024 were downloaded in May 2025. Entries in the dataset correspond to people who are reported by a clinician or epidemiologist as a “suspected” case, who then either have a laboratory test using ELISA (with IgM positive/negative results reported), are epidemiologically linked to another confirmed or suspected measles case or are not further followed up and considered “clinically compatible” (Appendix A, Figure S2). Excluding negative laboratory tests, between 2011 and 2024 there were 121,510 measles entries in the dataset (range 1,597-29,808 across years, Appendix A, Figure S1), of which 33.7% were laboratory tested, 64.3% were epidemiologically linked or clinically confirmed and 2% still had pending results (Appendix A, Figure S5). Of the 53,903 total laboratory tested cases, 34% were positive.

### 2.3 Data processing

For the purposes of this analysis the surveillance data were processed to produce a set of distinct outbreaks at the woreda level based on the Ethiopian definition of an outbreak: three laboratory-confirmed diagnoses in a woreda within 30 days, with the outbreak lasting until there were no new diagnoses (epidemiologically linked) for 42 consecutive days [4]. For each outbreak identified in the data, information was extracted for outbreak location, duration, and cumulative identified cases (laboratory diagnosed, epidemiologically linked or clinically compatible) by age (<18 years versus ≥18 years). Between 2011 and 2024 there were 2176 outbreaks identified across 679 woredas (Appendix A, Figure S6), with a median of 8 cases (IQR: 5-24) and a median duration of 81-days (IQR: 54-130-days). Further details are in Appendix A.

### 2.4 Model overview

A computational model was used to project cumulative measles infections and diagnoses over a 10-year period at a national level. This was done by simulating frequent smaller outbreaks at the woreda level and aggregating outcomes. By estimating national outcomes as the sum of outbreaks on small geographical areas, the model can capture the stochastic nature of outbreaks as well as the potential benefits of interventions to improve outbreak detection and response time by local public health teams.

### 2.5 Simulating individual outbreaks

#### 2.5.1 Outbreak model

An established agent-based model, Starsim [15], was used to simulate measles outbreaks in different Ethiopian woredas. The framework has previously been used to simulate outbreaks of measles and other pathogens in a variety of settings [16].

For each simulated outbreak, the model was initialised with a population of 50,000 agents with age distribution based on 2023 Ethiopian estimates [17] (Appendix B, Figure S11). Agents in the model were classified as susceptible, exposed, infectious or recovered following accepted measles modelling frameworks (Appendix B, Figure S10) [18]. The model includes three contact networks: household, school, and social networks. Household sizes and age distributions were based on Ethiopian household structures [19, 20]. Schools were implemented as collections of fully connected classroom clusters with a teacher assigned to each classroom, and additional random mixing between teachers and students in different classes within the same school. The social network was implemented as two random networks, separated for children aged 0-4 years and adults aged 19+ years (with schools approximating social mixing among people aged 5-18 years), and used to represent both social contacts and other daily contacts that could present opportunities for transmission. Contact networks were parametrized based off Ethiopian demographic data (Appendix B).

#### 2.5.2 Testing

Three different testing types were considered for this analysis: laboratory, epidemiological linkage and RDT (Appendix B, Figure S10). Each test type had parameters for testing probability (probability that a person with measles is tested using the test type), delay from test to result, sensitivity, specificity and unit cost (Table 1). Laboratory testing is assumed to be performed using ELISA. Testing probabilities for laboratory and epidemiological linkage reflect testing rates before and after outbreak detection respectively, and were both estimated through calibration (see below). Testing probability for non-measles cases that seek care were estimated using total negative tests from the data (Appendix A, Figure S3, Figure S4). Testing probabilities for RDTs were scenario-based. Delays for test results were based on the surveillance data for laboratory testing (median 6 days, range 1-332 days, Appendix A, Figure S5) and assumed to be zero for epidemiological linkage and RDTs. Sensitivity and specificity estimates were taken from the literature and unit costs were based on data from the Ethiopian Public Health Institute (Table 1, Appendix C).

**Table 1:** Model inputs. All costs are in 2025 USD.

| Parameter | Value | Source |
| --- | --- | --- |
| **Outbreak model** |  |  |
| **Immunity parameters** |  |  |
| Whole population initial immunity | 80% | Includes estimates of natural infection and vaccine acquired immunity, Trentini et al, 2017 [27] |
| Percentage immune population 9-months-4-years | 7% | Includes estimates of natural infection and vaccine acquired immunity, Trentini et al, 2017 [27] |
| Percentage immune population 5-17-years | 28% | Includes estimates of natural infection and vaccine acquired immunity, Trentini et al, 2017 [27] |
| Percentage immune population 18+ years | 65% | Includes estimates of natural infection and vaccine acquired immunity, Trentini et al, 2017 [27] |
| Protection from infection: single dose vaccination | 83% | Estimate for those ages 9-months and over. Lochlainn et al, 2019 [28] |
| Protection from infection: two dose vaccination or prior infection | 95% | Systematic review and meta-analysis. Hughes et al, 2020 [29] |
| **Disease severity** |  |  |
| Probability death given infection: <1 years old | 2.45% | Modelling study estimating measles case fatality ratios in LMICs. Sbarra et al, 2023 [24] |
| Probability death given infection: 1-4-years old | 1.17% | Modelling study estimating measles case fatality ratios in LMICs. Sbarra et al, 2023 [24] |
| Probability death given infection: 5-9-years old | 0.65% | Modelling study estimating measles case fatality ratios in LMICs. Sbarra et al, 2023 [24] |
| Probability death given infection: 10-14-years old | 0.5% | Modelling study estimating measles case fatality ratios in LMICs. Sbarra et al, 2023 [24] |
| Probability death given infection: | 0.26% | Modelling study estimating measles case fatality |
| 15+ years old |  | rations in LMICs. Sbarra et al, 2023 [24] |
| School network |  |  |
| Mean school size | 994 | Ethiopian annual education statistics, 2013 [30] |
| Mean class size | 50 | Ethiopian school survey. Aurino et al, 2012-2013 [31] |
| Primary school enrolment | 85% | Statistics on Ethiopian primary school enrolment, 2023 [32] |
| Secondary school enrolment | 30% | Profile on Ethiopian education, 2018 [33] |
| Social network |  |  |
| Mean social contacts | 6 | Estimates of contact patterns in Ethiopia. Trentini et al, 2023 [34] |
| Testing |  |  |
| Laboratory test: sensitivity | 0.97 | Systematic review of ELISA accuracy. Zubach et al, 2024 [9] |
| Laboratory test: specificity | 0.96 | Systematic review of ELISA accuracy. Zubach et al, 2024 [9] |
| Laboratory testing probability | 6% | Calibration, Section 2.5.4 |
| Epidemiological linkage: sensitivity | 0.907 | Observation diagnostic study in Indonesia. Husada et al, 2020 [35] |
| Epidemiological linkage: specificity | 0.286 | Observation diagnostic study in Indonesia. Husada et al, 2020 [35] |
| Epidemiological linkage probability | 10% | Calibration, Section 2.5.4 |
| RDT: sensitivity | 0.90 | Estimated using sampled collected from multi country surveillance programs. Rachlin et al, 2024 [36] |
| RDT: specificity | 0.95 | Estimated using sampled collected from multi country surveillance programs. Rachlin et al, 2024 [36] |
| Accelerated vaccine campaign |  |  |
| Final coverage 6-months-10 years old | 95% | Reactive immunisation campaign in 2024 over 10-days [22] |
| Time from outbreak detection to implementation | 8-10 weeks | Personal communication with EPHI [21] |
| Proportion outbreaks with vaccine response | 35% | Personal communication with EPHI [21] |
| Cost model |  |  |
| Surveillance costs |  |  |
| Laboratory test | \$160.47 | Include travel, staff and kit costs. Personal communication with EPHI [21] |
| Epidemiological linkage | \$84.5 | Includes travel and staff costs. Personal communication with EPHI [21] |
| RDT | \$4.29 | Includes shipment, staff and kit costs. Personal communication with EPHI [21] |
| Additional costs |  |  |
| Hospitalised measles case | \$22.79 | Cost-effectiveness analysis of Ethiopian measles outbreak, cost converted to US\$ 2025. Driessen et al, 2015 [23] |
| Daily outbreak costs | \$331.47 | Personal communication with EPHI [21] |
| Daily outbreak costs (including reactive vaccination) | \$497.32 | Personal communication with EPHI [21] |
| Additional inputs |  |  |
| Probability of measles case being hospitalised | 12.4% | Modelling study based on Ethiopian line listed measles data. Poletti et al, 2018 [37] |
| Discounting for disability-adjusted life years | 3% | Discounting applied annually. |
| Discounting for costs | 3% | Discounting applied annually. |
| Disability weight (severe disease) | 0.13 | Global burden of disease study, 2021 [26] |
| Disability weight (mild disease) | 0.05 | Global burden of disease study, 2021 [26] |

#### 2.5.3 Responses to outbreaks

Once an outbreak is detected in a woreda, a vaccine response may be initiated. Due to limited stocks and logistical constraints this is not always possible and is estimated to occur approximately 35% of the time, taking eight to ten weeks to initiate [21]. In the model, this was implemented by having an outbreak response immunisation intervention triggered with 35% probability once an outbreak is detected, taking nine weeks to commence, and achieving 95% coverage among children six months to ten years over an ~8-day period [22]. For the model population of 50,000 agents, this is estimated as vaccines delivered at a rate of ~500 people per day.

#### 2.5.4 Calibration

The model was calibrated such that when 5,000 outbreaks were simulated, the distribution of model outcomes for detected cases per outbreak, outbreak duration and the age distribution of detected cases aligned with the distribution of these measures within the processed outbreak data. This was achieved by fitting parameters for the laboratory testing probability, epidemiological linkage testing probability, and force of infection (probability of transmission per day per contact with an infectious person), since these parameters have the greatest uncertainty. A detailed description of the model calibration method is in Appendix B (Figure S12).

### 2.6 Projecting national 10-year outcomes

A baseline scenario was constructed where national 10-year measles burden was estimated assuming that the frequency of woreda-level outbreaks observed between 2011–2024 (~152 per year) would continue for each future year (Appendix A.2, Figure S7). A duration of 10-years was chosen to produce sufficiently stable outcomes but avoid uncertainty in potential long term demographic changes. A distribution of the number of outbreaks which occur per month at the woreda-level in Ethiopia was generated from the processed line-listed case data. This distribution was sampled each month over the 10-year period being considered. Additional weighting of outbreak probability by immunity levels across woredas were explored, however data inconsistencies meant this was not feasible (Appendix A.3). For each month of the projection, Ethiopia would experience N outbreaks, with each outbreak being assigned randomly to a woreda. If a woreda experienced an outbreak, a set of epidemiological and response outcomes (e.g., cases, deaths, vaccines, etc.) from a precomputed outbreak simulation produced by the measles outbreak model was sampled and scaled to the size of the woreda. A more detailed description of the method is in Appendix B.

The cumulative outcomes over 10-years were estimated by aggregating outcomes across all outbreaks for number of infections, diagnoses, deaths, tests used (by test type), days spent in a detected outbreak, and vaccine responses triggered.

### 2.7 Scenarios

Scenarios were chosen to assess the impact of scaling up RDT usage to enable early detection of outbreaks and faster case detection throughout outbreaks. Scenarios are tested as follows:

1. **S0: Current standards (baseline)**. Until an outbreak is detected laboratory testing only. After outbreak detection all diagnosis through epidemiological linkage.
2. **S1: Replace labs with RDTs**. Until an outbreak is detected RDTs only (using laboratory testing probability). After outbreak detection all diagnosis through epidemiological linkage.
3. **S2: Replace lab & epi with RDTs**. Until an outbreak is detected RDTs only (using laboratory testing probability). After outbreak detection RDTs only (using epidemiological linkage testing probability).
4. **S3a: Case detection rate doubled**. RDTs used to increase testing such that case detection rate doubles compared to the baseline scenario. Laboratory testing and epidemiological linkage are also replaced with RDTs.
5. **S3b: 100% case detection**. An optimistic upper bound scenario, where RDTs used to increase testing such that 100% case detection occurs. Laboratory testing and epidemiological linkage are also replaced with RDTs.

### 2.8 Costs

Total national costs of testing, outbreak response and hospitalisation of severe measles cases were calculated from a government perspective for each scenario for a 10-year projection period. Costs are presented in 2025 US$ with future costs discounted at 3% per annum.

Costs of laboratory testing, epidemiological linkage and daily outbreak response teams (including additional immunisation where it occurred) were obtained from Ethiopian Public Health Institute and included commodities, human resources, logistics and transport, active surveillance, cold chain and waste management and other reporting costs (Table 1 and Appendix C). Hospitalisation costs for severe measles cases were obtained from the literature [23]. RDT unit costs were estimated based on procurement costs and assumed similar delivery costs to other testing options. As the scenarios considering expanded RDTs are based on the probability of people with measles being diagnosed, the RDT positivity rate (and hence number of negative RDTs) is unknown. Therefore, in these scenarios we estimate the RDT positivity rate threshold that would still lead to scenarios being cost-effective.

### 2.9 Cost-effectiveness analysis

The health economic analysis follows the Consolidated Health Economic Evaluation Reporting Standards (CHEERS) (Appendix E), with a protocol included as a sub-component of the overall pilot RDT deployment study protocol. For each scenario total measles-related disability-adjusted life years (DALYs) were calculated. Years of life lost were calculated based on model outcomes for age-specific measles infections (as opposed to diagnoses), age-specific measles mortality rates [24] and estimated years of life remaining by age [25]. Years lived with disability were calculated based on disutility weights for severe and mild disease [26]. Future DALYs were discounted at 3% per annum.

For each scenario the difference in total costs and total DALYs were calculated compared to the baseline.

### 2.10 Uncertainty

For each scenario 100 projections of 10-year outcomes were made. For each projection, the number of outbreaks in each month of the 10-year period was sampled from the distribution of outbreaks per month in the data (Appendix A, Figure S7), and for each outbreak a stochastic model simulation was used which included sampling of calibration parameters.

For individual outbreaks total diagnosis, false positives, days to detect an outbreak, and outbreak duration are recorded. Over 10-years total DALYs, test costs, outbreak response costs, and hospital costs are recorded. Results are presented as the median and inter-quartile range of these model runs.

### 2.11 Patient and Public reporting

No patients were involved in this study.

## 3 Results

### 3.1 Model calibration

In the current standards (baseline) scenario, woreda-level outbreaks were estimated to have a median 9 (5–18) detected cases per outbreak and last a median 107 (65–170) days. Over 10 years 1468 (1420–1545) outbreaks were estimated to occur, giving a median 58 (55–62) thousand diagnoses and 59 (49–72) thousand DALYs (Figure 2, **Error! Reference source not found**., Figure S15). Through the model calibration process, due to the very low daily testing probability, it was estimated to take a median 113 (IQR 80–150) days from the introduction of a measles case to detection of an outbreak.

**Figure 1:**
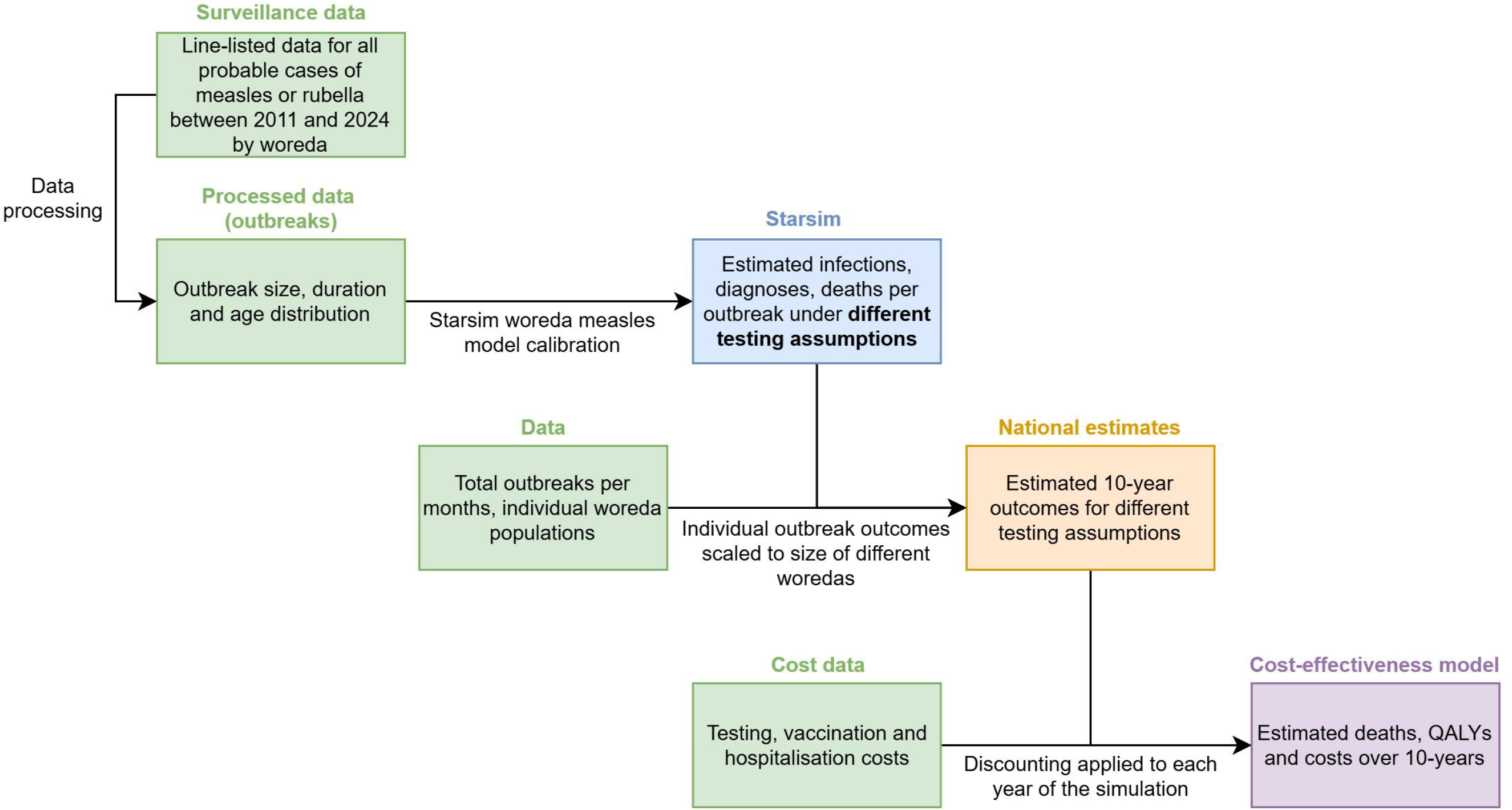
Modelling framework

**Figure 2:**
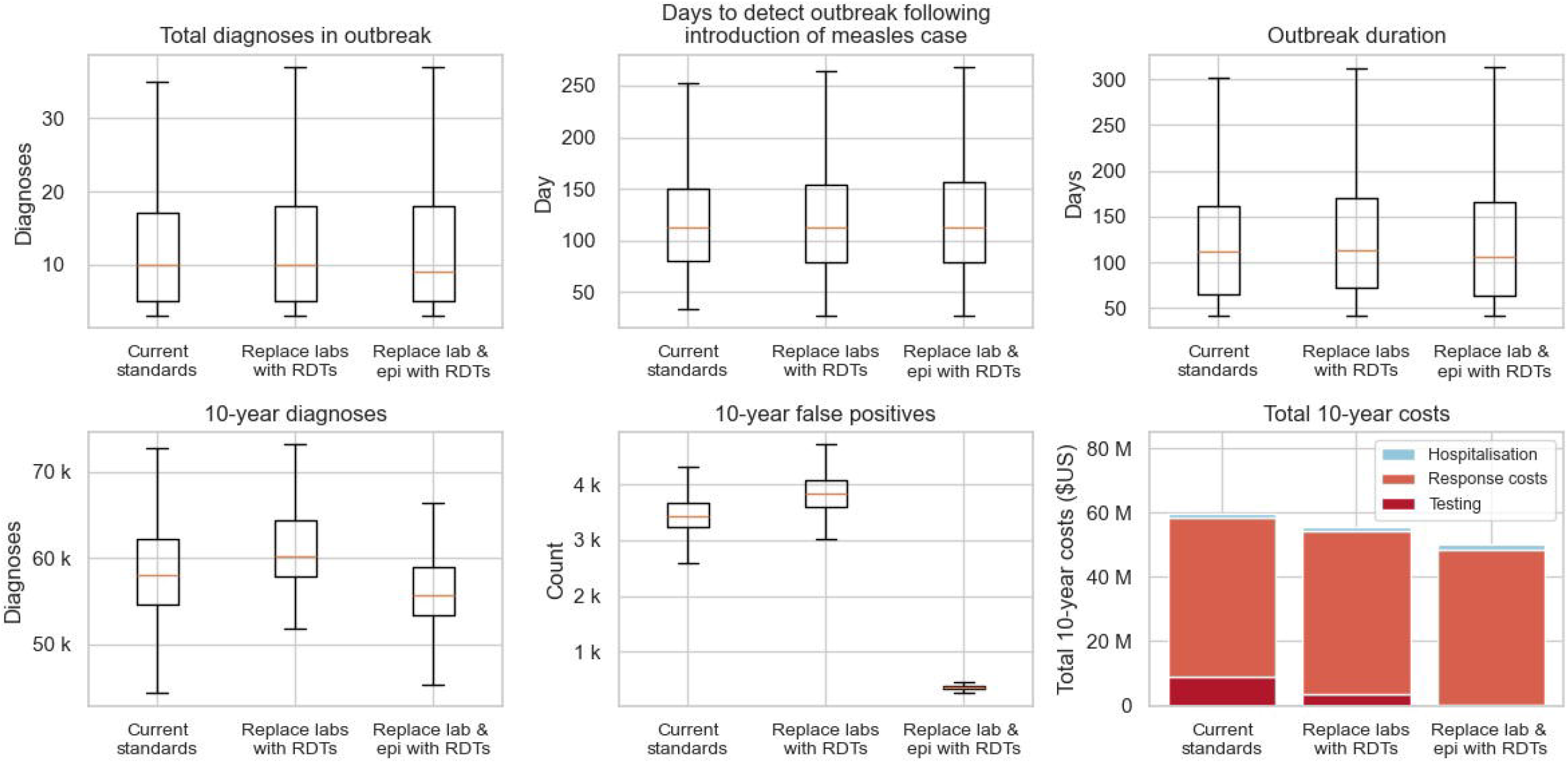
Outcomes per outbreak (top row) and over 10 years (bottom row) when using rapid diagnostic test to replace laboratory testing (“replace labs with RDTs”) or laboratory testing and epidemiological linkage (“replace lab & epi with RDTs”). Boxplots represent distribution of outcomes over simulations per scenario; the boxes cover the interquartile range, and the whiskers extend to the minimum and maximum value while excluding outliers (defined as simulations further than 2.5 times the inter-quartile range from the median).

Total difference in epidemiological outcomes between scenarios can be found in Appendix C, Table S4. The total 10-year cost of the baseline scenario was US$60 (US$56–64) million, of which 14.6% was for testing, 82.9% for outbreak response and 2.5% for hospitalisation.

### 3.2 RDT replacement scenarios

Compared to the baseline, all RDT scenarios were cost saving over a 10-year period, predominantly due to the high costs of laboratory testing and epidemiological linkage averted when switching to RDTs (Figure 2).

Replacing laboratory testing with RDTs gave similar outcomes for outbreak detection time (Figure 2). While RDTs eliminated the delay in receiving laboratory testing results, if they were only used at such low testing rates (equivalent to current laboratory) then the lower sensitivity of the RDTs (sensitivity 0.9, specificity 0.95) meant that the occasional missed cases negated the benefits of timely results with respect to outbreak size and duration. However, despite similar cumulative diagnoses and DALYs, replacing laboratory testing with RDTs saved US$4.2 million over the 10-year timeframe.

Replacing epidemiological linkage with RDTs, in addition to replacing laboratory testing with RDTs, resulted in a large reduction in false positive diagnoses due to the superior specificity of RDTs compared to epidemiological linkage. DALYs were also similar to the baseline, but due to shorter outbreak duration there were cumulatively fewer days requiring an outbreak response, and subsequently US$9.7 million was saved over 10 years compared to the baseline.

#### 3.4 RDT scale-up

Scenarios that used RDTs to increase testing rates, in addition to replacing laboratory testing and epidemiological linkage, are shown in Figure 3 and Table 2. Compared to the baseline, these scenarios reduced the time to outbreak detection and response, the number of false positives, the time spent in outbreaks and total costs.

**Table 2:** Differences in DALYs and costs per scenario compared to the baseline, shown as median and interquartile range.

| Baseline | Total DALYs | Total test costs (million US\$) | Total daily outbreak response costs (million US\$) | Total hospital costs (million US\$) | Total costs (million US\$) |
| --- | --- | --- | --- | --- | --- |
| Current standards | 59,360<br>(56,068, 64,650) | \$8.8<br>(\$8.4, \$9.1) | \$49.9<br>(\$46.2, \$52.9) | \$1.4<br>(\$1.3, \$1.5) | \$60.1<br>(\$55.9, \$63.5) |
| Scenario | Difference in total DALYs | Difference in total test costs (million US\$) | Difference in total daily outbreak response costs (million US\$) | Difference in total hospital costs (million US\$) | Difference in total costs (million US\$) |
| Current standards | Reference | Reference | Reference | Reference | Reference |
| Replace labs with RDTs | 2,632<br>(205, 5,660) | -\$5.1<br>(-\$5.3, -\$5.0) | \$1.0<br>(\$0.4, \$1.7) | \$0.0<br>(-\$0.0, \$0.1) | -\$4.2<br>(-\$4.9, -\$3.2) |
| Replace lab & epi with RDTs | 146<br>(-2,490, 2,719) | -\$8.4<br>(-\$8.8, -\$8.0) | -\$1.4<br>(-\$2.0, -\$0.7) | \$0.0<br>(-\$0.0, \$0.1) | -\$9.7<br>(-\$10.4, -\$9.0) |
| Replace lab & epi with RDTs, case detection rate doubled | -17,471<br>(-19,837, -15,598) | -\$8.4<br>(-\$8.7, -\$8.0) | \$4.4<br>(\$3.8, \$5.2) | -\$0.4<br>(-\$0.5, -\$0.4) | -\$4.3<br>(-\$5.0, -\$3.5) |
| Replace lab & epi with RDTs, 100% case detection | -50,620<br>(-55,464, -48,100) | -\$8.3<br>(-\$8.7, -\$7.9) | -\$9.7<br>(-\$10.8, -\$8.7) | -\$1.2<br>(-\$1.3, -\$1.1) | -\$19.1<br>(-\$20.6, -\$18.0) |

**Figure 3:**
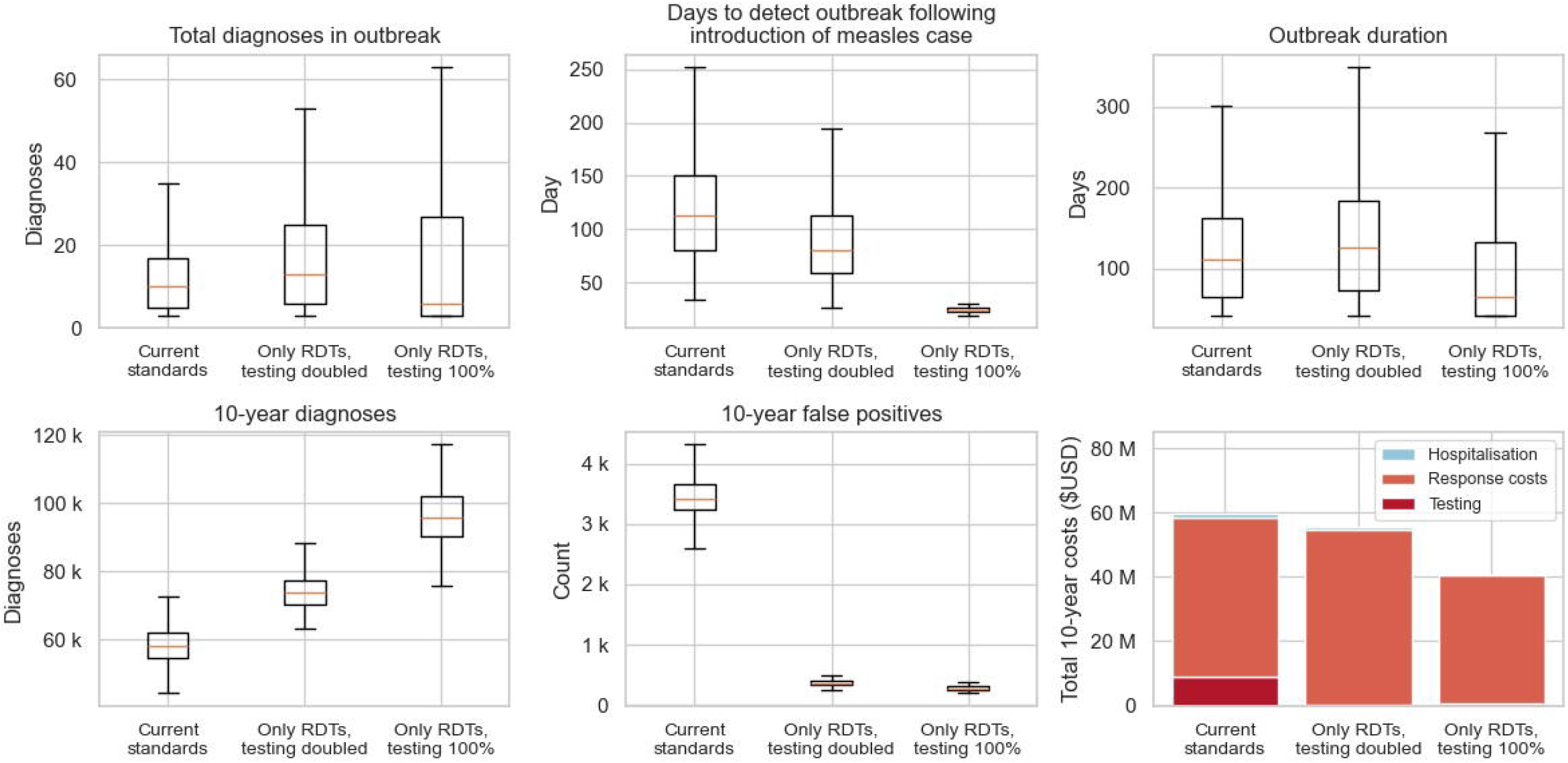
Outcomes per outbreak (top row) and over 10 years (bottom row) when using rapid diagnostic tests to expand testing coverage, in addition to replacing existing laboratory testing and epidemiological linkage. Boxplots represent distribution of outcomes over simulations per scenario; the boxes cover the interquartile range, and the whiskers extend to the minimum and maximum value while excluding outliers (defined as simulations further than 2.5 times the inter-quartile range from the median).

When the measles case detection rate was doubled there were a median of 12 (6–25) diagnoses per outbreak, an increase from the baseline due to detection of cases that were previously missed; however, due to faster outbreak detection (median of 80 (59–113) days) and response, there were fewer infections (Appendix D, Figure S13) and 17 (11–23) thousand DALYs averted compared to the baseline (Appendix D, Table S3, Figure S14). There was also US$4.3 million saved compared to the baseline, since even with the additional testing, total RDT costs were offset by savings from no longer undertaking laboratory tests or epidemiological linkage. These cost savings persisted in the model as long as RDT positivity rate was greater than 6.02% (5.25–7.35%).

Similar results were observed for the optimistic 100% detection rate scenario using RDTs (i.e. faster outbreak detection and response, more diagnoses but fewer infections and DALYs and costs), with the exception that the median diagnoses per outbreak decreased compared to the baseline, due to detection of smaller outbreaks that would otherwise have been missed.

## 4 Discussion

To effectively control and respond measles outbreaks in low- and middle-income countries such as Ethiopia, timely and cost-effective detection of cases is critical. This analysis used an agent-based model to simulate measles outbreaks in Ethiopian woredas and assess the cost-effectiveness of introducing RDTs for enhanced measles surveillance. The study found that using RDTs to replace current laboratory testing and epidemiological linkage with or without expanded testing is likely to be cost saving, but health outcomes of scenarios varied. Replacing laboratory testing with RDTs without also expanding testing coverage had minimal impact on outbreak detection times and DALYs, because the benefits of eliminating delays in laboratory results were offset by missed cases from the lower RDT sensitivity. However, when RDTs were used to additionally expand testing coverage, the lower sensitivity was less of an issue, and outbreak detection and response times were reduced leading to fewer infections and DALYs. These results generally support the widespread use of RDTs from a health economics perspective where laboratory testing capacity is limited.

Replacing laboratory testing with RDTs without expanding testing could save Ethiopia US$4.2 million over a 10-year period. Measles laboratory testing was introduced to aid surveillance in Ethiopia in 2003 and is currently the only diagnostic tool for detecting and declaring measles outbreaks [12]. While the cost of a single laboratory kit is relatively cheap (US$10.86), the entire process from specimen collection to result has multiple procedures which compound costs. Ethiopia currently has very few operational laboratory testing facilities. As a result, all laboratory testing incurs high travel costs (both transportation and staff) as well as laboratory staff salaries, and when these factors are considered, the estimated cost is US$160.47 in total per test (Appendix C). By comparison, RDTs can substantively reduce travel costs and limit the number of staff involved in a single diagnosis. Hence, their estimated unit cost is approximately forty times cheaper than laboratory testing (US$4.29). While there is uncertainty around the exact unit costs of RDTs since they are not yet commercially available, it is very likely that in the Ethiopian context they would remain an order of magnitude cheaper than laboratory testing.

Reducing the time to detect and respond to outbreaks can improve outcomes [16]. In this analysis there was a trade-off observed when RDTs were used to replace laboratory testing: RDTs reduced outbreak detection time by eliminating the median six-day delay to obtain laboratory test results, but in some model simulations increased outbreak detection time when the slightly lower RDT sensitivity (90% compared to 97% for laboratory testing) led to additional cases being missed. This was particularly relevant because the definition of an outbreak used by public health teams is only three cases. However, the model also suggests that the lower test sensitivity can be offset by increased testing, and in scenarios where RDTs were used to expand testing as well as replace current laboratory testing, faster outbreak detection times were observed with corresponding health benefits.

Currently in Ethiopia once an outbreak is detected epidemiological linkage is used in place of laboratory testing for case detection. Epidemiological linkage is relatively resource intensive with a high cost per case detected (US$84.5, Appendix C), since it requires a team of staff and significant travel to contact trace suspected measles cases. Epidemiological linkage also has a low specificity (estimated 28.6% [35]) due to similarities in clinical presentation among people with measles and non-measles febrile illnesses. The low specificity can lead to false positive measles diagnoses during an outbreak, which can delay when the outbreak is declared over (i.e. having no new diagnoses for 42 consecutive days). When RDTs were modelled to replace epidemiological linkage post-outbreak detection, we found that testing costs were substantively reduced since RDTs could be administered by lower cadre healthcare workers and fewer resources were required for contact tracing, and also that total daily outbreak response costs were reduced because fewer false positive diagnoses meant a shorter outbreak duration. In the model, replacing epidemiological linkage with RDTs had no impact on DALYs, because infections were only influenced by the speed of response (determined by outbreak detection day), rather than testing methods after the outbreak was detected.

Some consideration is needed for how test positivity rates may change with increased RDT availability, and the impact this could have on total costs and cost-effectiveness. The scenarios where RDTs were used to simply replace current testing were found to be cost saving, based on test positivity rates and hence the cost of negative tests derived from the data. However, increased RDT availability may result in lower thresholds for when health care workers decide to conduct tests. It is unknown how much this would occur when expanding testing, but the model results suggest that there is a significant scope for having lower test positivity before it would no longer be cost saving to expand coverage. For example, in the scenario where the probability of a measles case being diagnosed was doubled, test positivity could reduce to as low as 6.02% before it was no longer cost saving compared to the baseline (which had a test positivity rate of 34%).

There are limitations to this analysis. First, limited laboratory testing capacity means that the surveillance data used for model calibration may not be representative of measles epidemiology in Ethiopia. The model was calibrated to detected cases, with total infections being a model outcome that was used to estimate DALYs for each scenario. Furthermore, because the daily probability of testing is treated as a single calibration parameter, the model does not capture variation in testing behaviour or testing capacity across woredas. Second, each woreda was assumed to have the same measles vaccine coverage (the national estimate), since administrative vaccine data at the woreda-level were inconsistent and unable to be incorporated in the analysis (Appendix A, Figure S8, Figure S9). The 10-year projections also did not account for immunity gained through reactive vaccination campaigns or infections during an outbreak, which may affect woredas that have repeat outbreaks. Together these immunity assumptions mean that model outcomes are more representative of national averages with limited capture of heterogeneity across the country. Third, because the 10-year projections are informed by historical outbreak frequency, they may not fully capture the any future changes in measles incursion rates or vaccination. Fourth, RDT characteristics (unit costs, sensitivity, specificity) are based on limited available data, and future studies may lead to different estimates for these parameters, particularly once RDTs become commercially available.

## 5 Conclusion

In Ethiopia, use of measles RDTs could be cost saving, and if used to expand rather than just replace current testing could prevent measles infections through faster outbreak detection and response.

## Data Availability

Data sharing: Model parameters are available in the supplementary material, and the model code is available online: https://github.com/Burnet-Modelling/eth_measles_outbreaks.

## Supporting information

### Funding

This study was funded by Gavi, the vaccine alliance (Contract Number GAVI100001110). Gavi contributed to study objectives, but had no role in study design, conduct, analysis and manuscript writing including the decision to publish the results.

## Acknowledgements

Health Poverty Action and Ethiopian Public Health Institute staff are acknowledged for coordination and management of data extraction and transferring. FM, FF and NS are the recipients of Australia National Health and Medical Research Council fellowships. Burnet Institute authors gratefully acknowledge the support provided by the Victorian Government Operational Infrastructure Support Program to Burnet Institute.

## Ethics statement

This study is the secondary data analysis and modelling of deidentified national case-based data and programmatic cost data. The study was approved by Alfred Ethics Committee, Australia (Project Number: 117210) and the Ethiopian Public Health Association Ethics Committee (EPHA/06/301/25).

## Medical writer or editor

There was no medical writer or editor involved in the creation of this manuscript.

## Manuscripts disputing published work

Not applicable.

## Related manuscripts

There is no manuscript, or any related manuscripts, currently under consideration or accepted elsewhere.

## Consent for publication

Not applicable.

## Data sharing

Model parameters are available in the supplementary material, and the model code is available online: https://github.com/Burnet-Modelling/eth_measles_outbreaks.

## Authors’ contributions

NS, WHO, FF and MM conceived and designed the study. BY, MM and MA extracted the data. KMK, KM, FM, FF and WHO managed and analysed the datasets. FM developed the model and ran the scenarios, with support from DD, RGA and NS. FM and NS wrote the first draft of the manuscript. DD, RGA, KYK, MS and RG contributed to manuscript drafting. All authors reviewed the manuscript, provided critical inputs and approved the final manuscript.

