## Supplementary material for "Cost-effectiveness of measles rapid diagnostic tests for replacing or expanding laboratory testing in Ethiopia"

Appendix A Data
           A.1 Measles surveillance data

Line listed measles case data was supplied between years 2011-2024. Each row in the dataset corresponded someone who had sought care and was suspected of either measles or rubella. Dataset included indicators such as age, case confirmation type (including negative laboratory tests), woreda, date onset of symptoms and laboratory delay from specimen collection to result.

Total negative tests were used to inform the rate of people who do not have measles but seek care (i.e. symptomatic for another infection but not measles) (Figure S3, Figure S4). This rate was estimated by disaggregating the total negative tests into years, then dividing by the total Ethiopian population in that year. For this analysis it was assumed that 2011 was an outlier due to it being the first recording year, hence is likely incomplete. The years 2020-2021 were also considered outliers due to those years intersecting with the COVID-19 period, hence testing rates for measles and rubella would likely be lower than previous years. Using these assumptions, it was estimated that the asymptomatic testing rate was 0.00125 to 0.00425% per year.

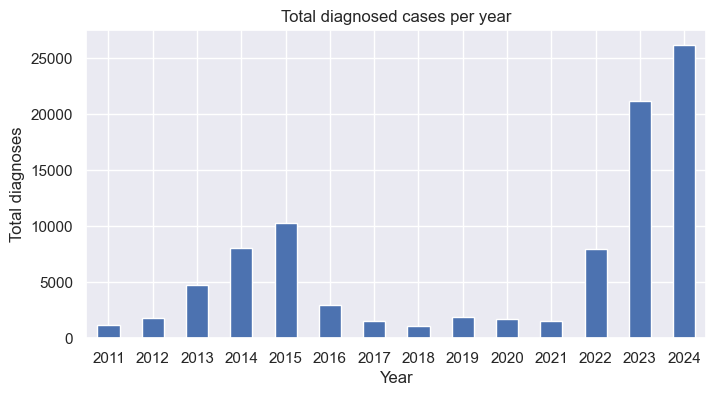

Figure S1: Total diagnosed measles cases per year between 2011-2024. Includes positive laboratory tests, epidemiologically linked cases, and clinically compatible cases.

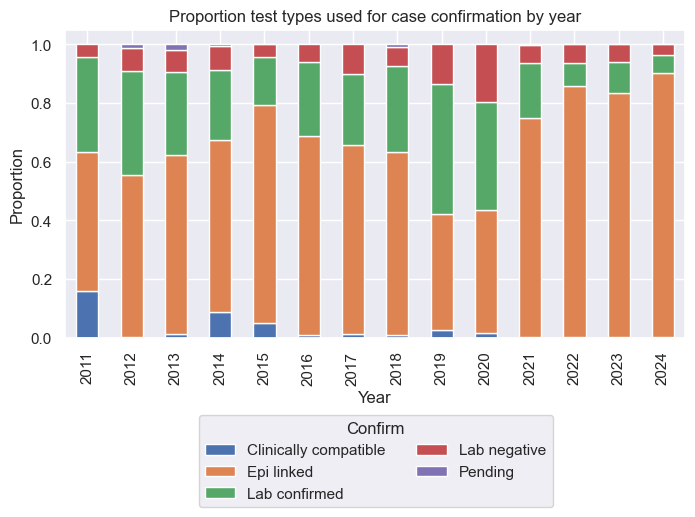

Figure S2: Distribution of test types by year. Laboratory tests have been disaggregrated into three categories: laboratory confirmed positive measles, laboratory confirmed negative measles and laboratory tests that are still pending results.

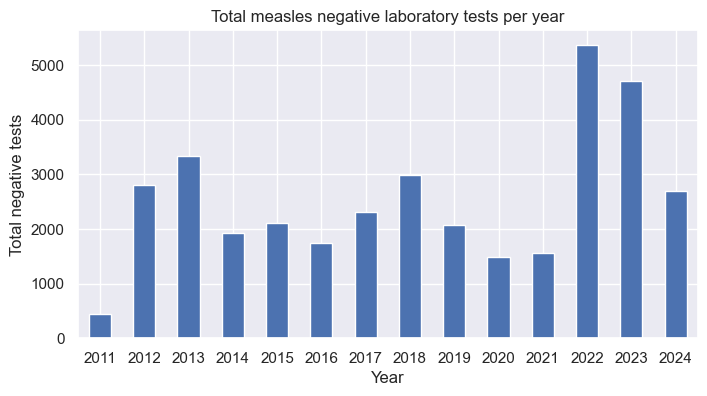

Figure S3: Total negative measles laboratory tests per year between 2011-2024.

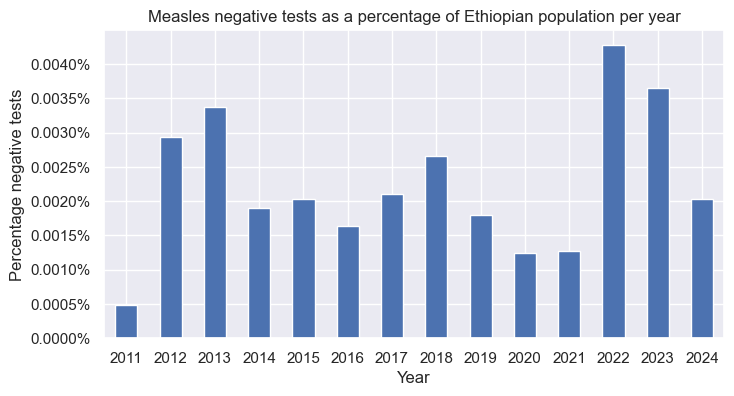

Figure S4: Percentage of negative laboratory tests for the whole Ethiopian population. Yearly negative tests have been divided by total Ethiopian population in that year. This is used as a proxy in the model for probability that someone without measles seeks care and is tested.

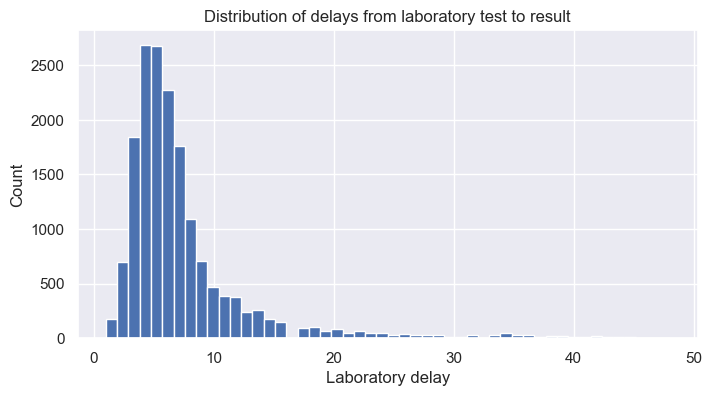

Figure S5: Distribution of laboratory delays. Data presents as approximately a negative binomial distribution with a mean of 7-days.

A.2 Data after processing into outbreaks

Line listed data was processed into individual outbreaks using the definition of: outbreak detected on three laboratory confirmed positive measles cases within a 30-day period, and outbreak ends given no new epidemiological linked cases in a 42-day period.

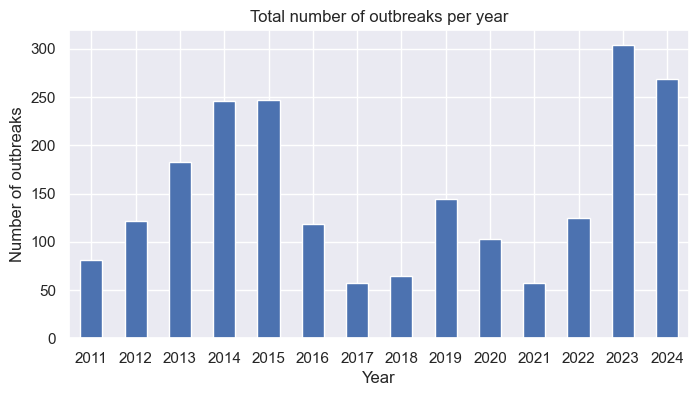

Figure S6: Total number of detected measles outbreaks (at least 3-laboratory confirmed cases within 30 days in the same woreda) per year between 2011-2024.

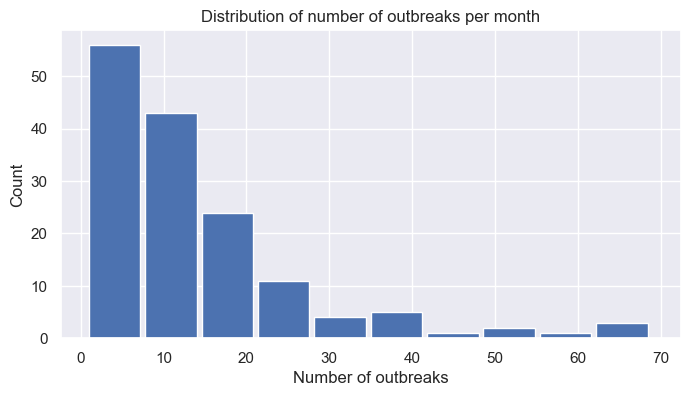

Figure S7: Distribution of the national number of woreda-level outbreaks per month, taken from all months between 2011-2024.

A.3 Vaccine data

For this analysis we assumed all outbreaks had the same vaccine coverage at the national estimates. Although administrative vaccination data disaggregated to the woreda level was available, it was excluded from the study as it was inconsistent and there was no observed relationship between vaccine coverage and detected measles cases (limiting calibration feasibility). For example, many of the woredas have a recorded measles vaccine coverage of over 100% (Figure S8). When the woreda-level vaccine coverage data was linked with the outbreak dataset, no consistent associations were observed between vaccine coverage and outbreak indicators such as measles diagnosis rate (Figure S8).

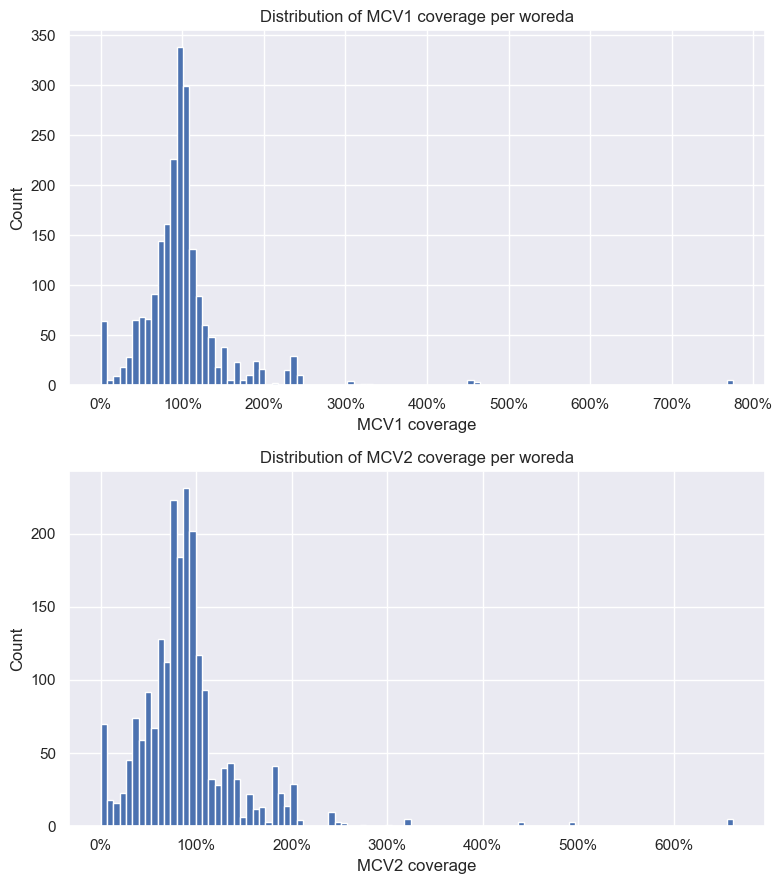

Figure S8: Distribution of woreda level MCV1 and MCV2 coverage for woredas in all detected outbreaks between 2011-2024.

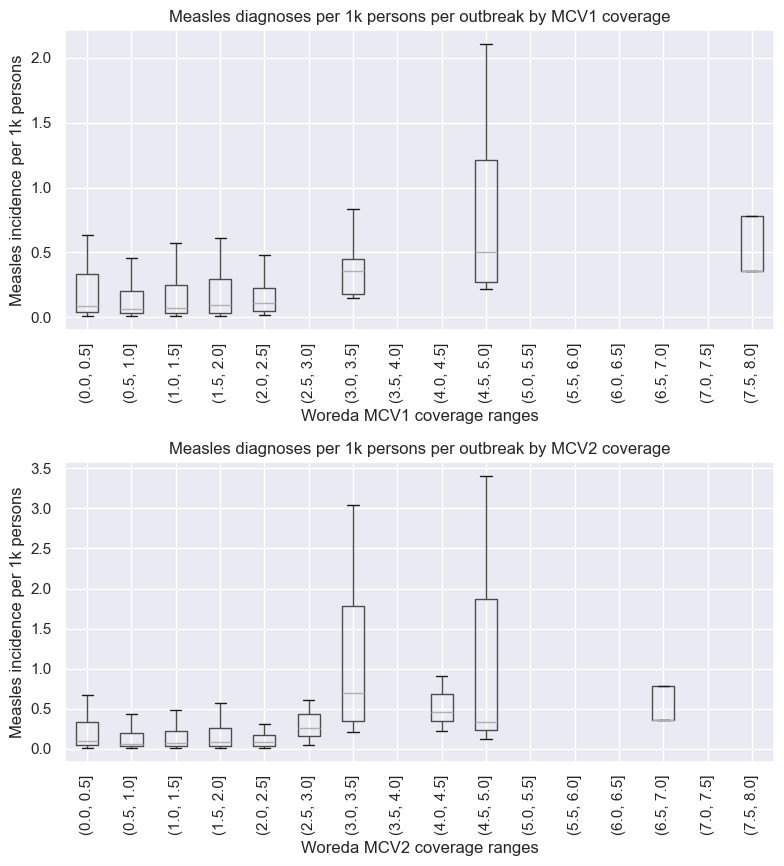

Figure S9: Distribution of measles diagnoses per 1k persons per outbreak for different woreda level values of first dose of measles-containing vaccine (MCV1) and second dose of measles-containing vaccine (MCV2) coverage

Appendix B Model details
           B.1 Model schematic

An agent-based model was used to simulate individual outbreaks. An SEIR (Susceptible, Exposed, Infected, Recovered) framework was used, which is the standard when modelling measles [1]. Each agent in the model has a daily probability of being tested or epidemiologically linked (Figure S10).

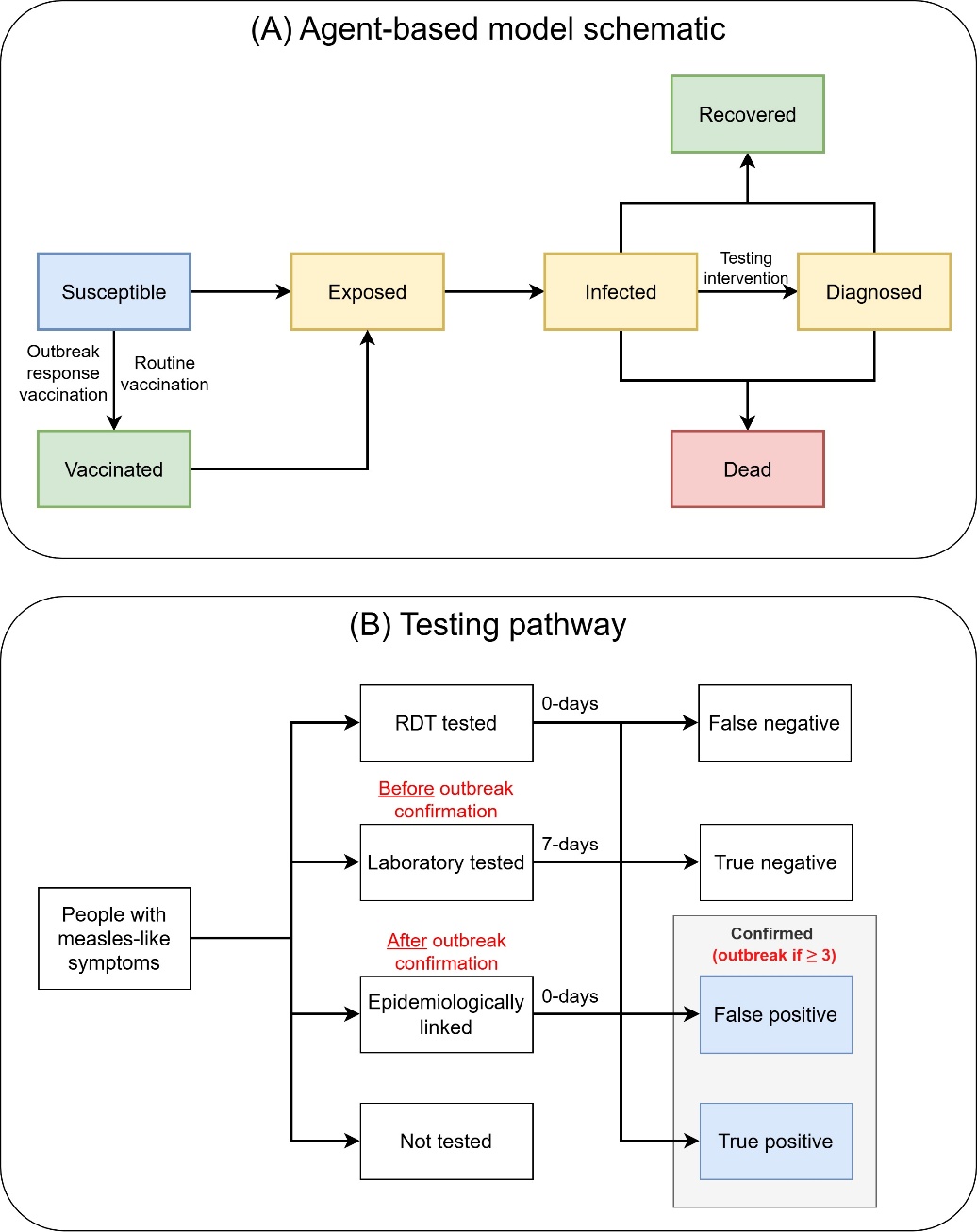

Figure S10: Model schematic. (A) model schematic displaying movement between disease states in agent-based model, (B) testing pathway for agents with measles-like symptoms.

B.2 Contact networks
    B.2.1 Household networks

A synthetic model population is initialized comprising of 50,000 agents. The age and household size structure of the model population is based on the Ethiopian population. Households are constructed by selecting a household head at random from the model population and a household size from the distribution of household sizes (Figure S11). The age of the household head is then used to inform the ages of the remaining household members, informed by the Ethiopia household mixing matrix from Prem et al [2].

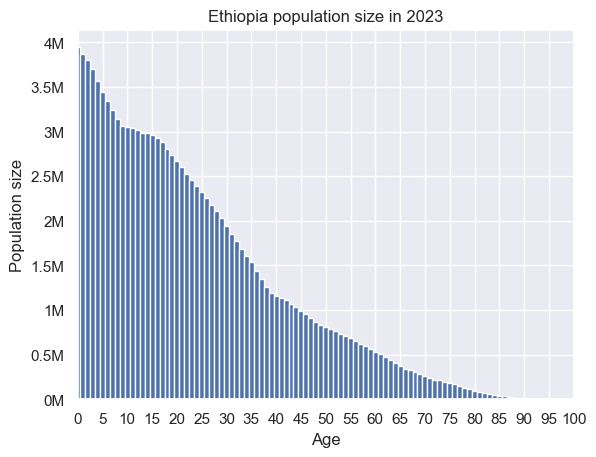

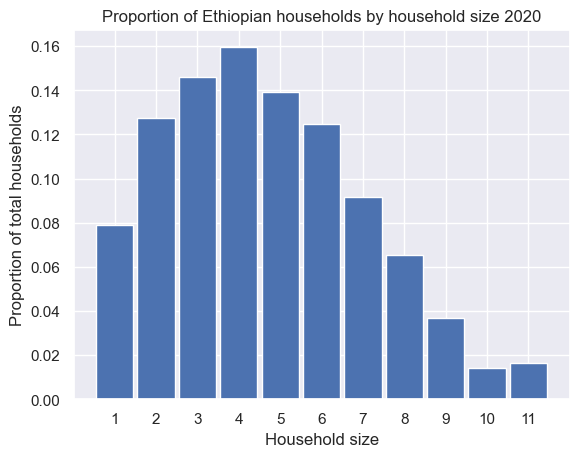

Figure S11: (Left) Age distribution of 2023 Ethiopian population. (Right) Distribution of Ethiopian household sizes in 2020.

B.2.2 Schools

Schools are modelled as a collection of classrooms that are aggregated into schools. Each student is assigned a classroom with others of the same age, and one teacher is assigned to each classroom drawn at random from the collection of agents 19 years or older. Average school and classroom sizes are based off Ethiopian data [3, 4]. Agents 7-18-years-old are assumed to be school aged. Primary school (ages 7-12) and secondary school (ages 13-18) have different enrolment rates based off Ethiopian school data [5, 6]. School mixing includes student-student contacts within classrooms, student-student contacts between students in different classrooms, teacher-teacher contacts and teacher-student contacts within classrooms they are assigned to.

B.2.3 Social

Two random networks were used to represent contacts outside of household and school settings (such as workplaces and social interactions). The two networks were separated by age, one for children aged 0-4 (not yet in school) and one for adults age 19 and above (no longer at school). Each agent was assigned a mean of 6-contacts with eligible agents within their respective random network [7]. The child social network assumed a higher beta due to children having a higher probability of transmitting infections between one another [8].

B.3 Model calibration

The model was calibrated such that when many outbreaks were simulated, the distribution of model outcomes for detected cases per outbreak, outbreak duration and the age distribution of detected cases aligned with the distribution of these measures within the processed outbreak data. This was achieved by fitting parameters for the laboratory testing probability, epidemiological linkage testing probability, and force of infection (probability of transmission per day per contact with an infectious person), since these parameters have the greatest uncertainty.

The algorithm to calibrate the model was: (1) sample testing probability and force of infection parameters from within their plausible ranges; (2) for each sampled parameter set, run an outbreak simulation; (3) classify the simulated outbreaks according to whether an outbreak was detected or not, and if an outbreak was detected whether the cumulative detected cases was within the inter-quartile range of the processed outbreak data; (4) selecting values for the sampled parameters based on a maximum likelihood for producing cumulative detected cases within the inter-quartile range of the processed data for simulations where an outbreak was detected.

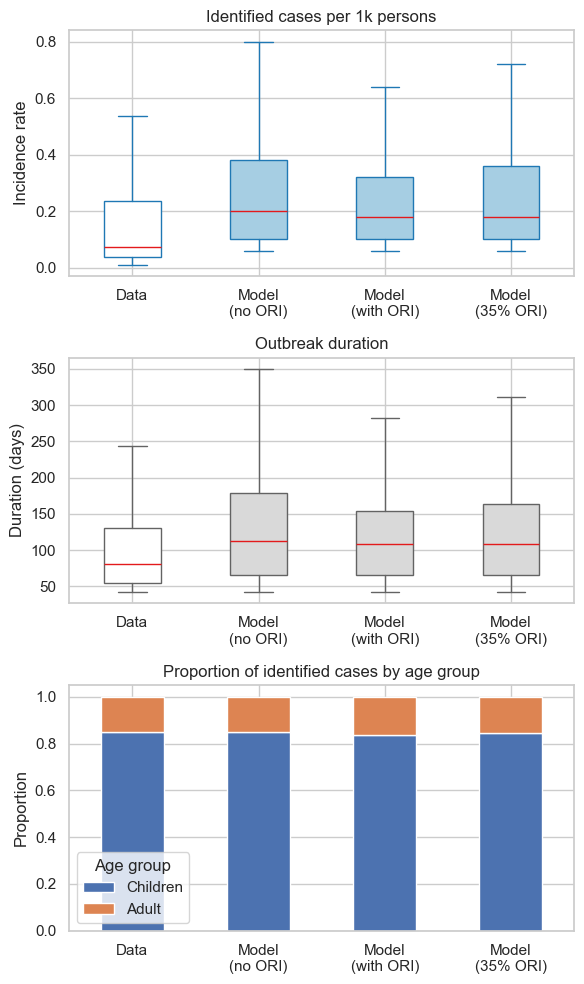

Figure S12: Model outcomes given no reactive vaccine campaign, 100% of the simulations have reactive vaccine campaigns and 35% of the simulations have reactive vaccine campaign. Model is fit to the distribution of diagnoses per 1k persons per outbreak, outbreak duration, and proportion of cases by age group (adults vs children).

B.4 10-year projections

To evaluate the impacts of different testing strategies on measles outbreaks over a 10-year time horizon, we developed an outbreak-tracking simulation for Ethiopia with monthly timesteps. The simulation takes estimates of country-level outbreak frequency and results from calibrated measles outbreak simulations as inputs, maps them to woredas, and synthesises them into a stochastic series of outbreaks.

B.4.1 Outbreak frequency estimates

A distribution of the number of outbreaks which occur per month at the woreda-level in Ethiopia was generated from the processed line-listed case data from 2011–2024. This distribution was sampled by the outbreak-tracking simulation during its monthly timestep to estimate the number of outbreaks which occurred each month during the simulation. When N > 0 outbreaks were determined to occur during a timestep, each outbreak was then assigned randomly to a woreda. The assigned woreda names were recorded and used to determine the at-risk population sizes for the outbreaks. When an outbreak was assigned to a woreda during this process, the woreda would be removed from the pool of potential outbreak locations until the outbreak was determined to have ended within the simulation. This was necessary because the simulation had a monthly timestep, however outbreaks typically last for longer than a single month, and we did not want to assign a ‘new’ outbreak to a woreda with an ongoing outbreak.

B.4.2 Outbreak size and response estimates

Using the calibrated measles outbreak model, sets of 5000 outbreak simulations were precomputed for each testing scenario being considered. Outcomes from these simulations were used to create an ‘outbreak library’, where modelled health and response outcomes from the sets of 5000 simulations were recorded. These outbreak simulation results were sampled again by the model, with the randomly selected outcomes representing a stochastic realisation of an outbreak in the woreda population assigned by the model. This sampling occurred during the monthly timestep of the outbreak-tracking simulation when N > 0 outbreaks were determined to occur, with the N sampled outcomes then scaled according to the population sizes of the assigned woreda for each outbreak.

B.4.3 Simulation aggregation and stochasticity

Cumulative outcomes for a single 10-year simulation were estimated by aggregating outcomes across all outbreaks for number of infections, diagnoses, deaths, tests used (by test type), days spent in a detected outbreak, and vaccine responses triggered. For each testing scenario, 100 simulations were run, each producing different cumulative outcomes due to variations in both the number of outbreaks sampled during each monthly timestep and the sampled outcomes for each outbreak.

Appendix C Costs

Daily costs associated with an outbreak and costs per testing type were supplied by EPHI in Ethiopia Birr (ETB) [9]. A conversion of 1 ETB to US$0.0065 in 2025 was used. Total daily costs during an outbreak included costs of training, surveillance, staff and transport (Table S1). For outbreaks that included a vaccination response this also included costs for vaccine and supplies, vaccinators and cold chain operations (Table S1). Estimated costs for laboratory confirmation, epidemiological linkage and RDTs included associated costs of kit, transport and staff (Table S2). The cost of RDTs is likely to change but is still expected to cost considerably less than laboratory confirmation and epidemiological linkage.

Table S1: Table of costs associated with a single day of an outbreak. Costs include daily costs of vaccines, staff, transport and supplies. Total daily cost of an outbreak displayed in 2025 ETB ans converted to 2025 USD for the main analysis.

| **Estimated Daily cost for Outbreak** | | | | | |
| --- | --- | --- | --- | --- | --- |
| **Category** | **Line Item** | **Unit Cost (ETB)** | **Qty/Day** | **Daily Cost (ETB)** | **Notes** |
| Vaccines & Supplies | Vaccine dose (incl. syringe, diluent, wastage) | 45 | 200 doses | 9,000 | Assuming 200 doses/day |
|  | Safety boxes | 120 | 5 | 600 | 1 per ~200 doses |
|  | Cold box/icepack operations | 1000 | 1 | 1000 | Fuel/electricity estimate |
| HR | Vaccinators (per diem) | 1500 | 10 | 15,000 | 10 vaccinators |
|  | Supervisors | 1500 | 2 | 3,000 | 2 supervisors |
|  | Data clerk | 1000 | 1 | 1000 |  |
|  | Drivers | 1500 | 2 | 3,000 | Vehicle + allowances |
| Training Cost | Team training (1-off cost) | — | — | 8,333 | Assuming one training per month per outbreak |
| Logistics & Transport | Fuel for outreach | 90/litre | 80 L | 7,200 | 2 vehicles doing 40 L/day |
|  | Vehicle rental/maintenance | 3,000 | 2 | 6,000 | 2 vehicles/day |
| Active Surveillance | Surveillance visits | 150/visit | 30 | 4,500 | CHW + transport |
|  | Lab sample transport | 200/sample | 5 | 1,000 | If samples daily |
| Social Mobilization / RCCE | Community mobilizers | 500 | 5 | 2,000 | Mobilizers/day |
|  | Radio spot (amortized/day) | — | — | 500 | 15,000 ETB ÷ 30 days |
| Cold Chain Ops | Generator fuel / electricity | 1500 | 1 | 1500 | Continuous cold chain |
| Waste Management | Sharps & waste disposal | 1000 | 1 | 1000 | Incineration/transport |
| Supplies & PPE | PPE & sanitation | 2500 | 1 | 2500 | Masks, gloves, sanitizers |
| M&E / Reporting | Data bundles & reporting | 500 | 1 | 500 | Daily coordination |
| Coordination & Overheads | Coordination cell costs | — | — | 1,500 | Meeting, utilities |
| Contingency | 10% of total | — | — | 7,632 |  |
|  |  | **Total daily costs (ETB)** | | 76,765 |  |
|  |  | **Total daily costs (US$)** | | $497.32 |  |
|  |  | **Total day costs without reactive vaccination (ETB)** | | 51,585 |  |
|  |  | **Total daily costs without reactive vaccination (US$)** | | $331.47 |  |

Table S2: Estimate unit costs in ETB for the three different testing types considered in the modelling: laboratory confirmation, epidemiological linkage and RDTs. Laboratory confirmation and epidemiological linkage include health worker and transport costs, and additional laboratory confirmation includes unit cost for ELISA kit. RDTs are not yet commercially available in Ethiopia, however the estimates below include health worker training and usage costs, and costs in transporting the tests to clinics.

|  | **Laboratory testing** | | | **Epidemiological linkage** | | | **RDT** | | |
| --- | --- | --- | --- | --- | --- | --- | --- | --- | --- |
| **Category** | **Line item** | **Value / Cost (ETB)** | **Unit cost (ETB)** | **Line item** | **Value / Cost (ETB)** | **Unit cost (ETB)** | **Line item** | **Value / Cost (ETB)** | **Unit cost (ETB)** |
| **Test kit** | Price of single ELISA (90 tests) | 98,000 | 1089 | NA | NA | NA | Single RDT | 462 | 462 |
|  | Additional cost of supplies | 609 | 609 |  |  |  |  |  |  |
| **Health workers** | Number of measles lab workers | 13 | 334 | ~5 health workers for 1.5 hours | 425 | 425 | Worker training and time to administer test | 20 | 20 |
|  | Total worker time spent on measles testing | 50% |  |  |  |  |  |  |  |
|  | Average staff monthly salart | 17,000 |  |  |  |  |  |  |  |
|  | Average tests per month | 331 |  |  |  |  |  |  |  |
| **Travel** | Air travel transport | 12,000 | 12,000 | Travel for active search and identification | 3,500 | 3,500 | Shipment and transport to healthcare facilities | 35 | 35 |
|  | Land travel transport | 5,000 | 5,000 |  |  |  |  |  |  |
|  | Staff per diem for transport | 4,250 | 4,250 |  |  |  |  |  |  |
| **Other costs** | Facility costs | 10,000 | 10,000 | Data clerk and communication | 9,156 | 9,156 | Reporting materials and communication | 150 | 150 |
|  | **Total unit cost (ETB)** | | 24,782 | **Total unit cost (ETB)** | | 13,081 | **Total unit cost (ETB)** | | 667 |
|  | **Total unit cost (US$)** | | $160.47 | **Total unit cost (US$)** | | $84.50 | **Total unit cost (US$)** | | $4.29 |

Appendix D Additional results

Table S3: Epidemiological outcomes per scenario, shown as median and inter-quartile range.

| **Scenario** | **Total diagnoses in outbreak** | **Days to detect outbreak following introduction of measles case** | **Outbreak duration** | **10-year diagnoses** | **10-year false positives** |
| --- | --- | --- | --- | --- | --- |
| Current standards | 9  (5, 18) | 113  (80, 150) | 107  (65, 170) | 57985  (54564, 62170) | 3423  (3228, 3663) |
| Replace labs with RDTs | 10  (5, 18) | 112  (79, 154) | 115  (72, 174) | 60254  (57818, 64426) | 3844  (3603, 4081) |
| Replace lab & epi with RDTs | 9  (5, 17) | 112  (79, 156) | 106  (64, 163) | 55747  (53425, 58924) | 350  (325, 379) |
| Replace lab & epi with RDTs, case detection rate doubled | 12  (6, 25) | 80  (59, 113) | 124  (74, 185) | 73743  (70314, 77460) | 368  (341, 408) |
| Replace lab & epi with RDTs, 100% case detection | 6  (3, 27) | 24  (23, 26) | 65  (42, 134) | 95892  (90165, 102156) | 281  (259, 310) |

Table S4: Difference in epidemiological outcomes per scenario, shown as median and inter-quartile range

| **Baseline** | **Total diagnoses in outbreak** | **Days to detect outbreak following introduction of measles case** | **Outbreak duration** | **10-year diagnoses** | **10-year false positives** |
| --- | --- | --- | --- | --- | --- |
| Current standards | 9  (5, 18) | 113  (80, 150) | 107  (65, 170) | 57985  (54564, 62170) | 3423  (3228, 3663) |
| **Scenario** | **Reduction in total diagnoses in outbreak** | **Reduction in days to detect outbreaks following introduction of measles case** | **Reduction in outbreak duration** | **Reduction in 10-year diagnoses** | **Reduction in 10-year false positives** |
| Current standards | Reference | Reference | Reference | Reference | Reference |
| Replace labs with RDTs | -1  (0, 0) | 1  (1, -4) | -8  (-7, -4) | -2269  (-3254, -2256) | -421  (-375, -418) |
| Replace lab & epi with RDTs | 0  (0, 1) | 1  (1, -6) | 1  (1, 7) | 2238  (1139, 3246) | 3073  (2903, 3284) |
| Replace lab & epi with RDTs, case detection rate doubled | -3  (-1, -7) | 33  (21, 37) | -17  (-9, -15) | -15758  (-15750, -15290) | 3055  (2887, 3255) |
| Replace lab & epi with RDTs, 100% case detection | 3  (2, -9) | 89  (57, 124) | 42  (23, 36) | -37907  (-35601, -39986) | 3142  (2969, 3353) |

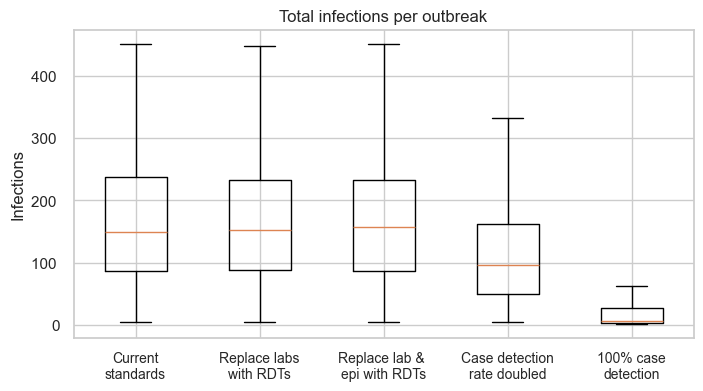

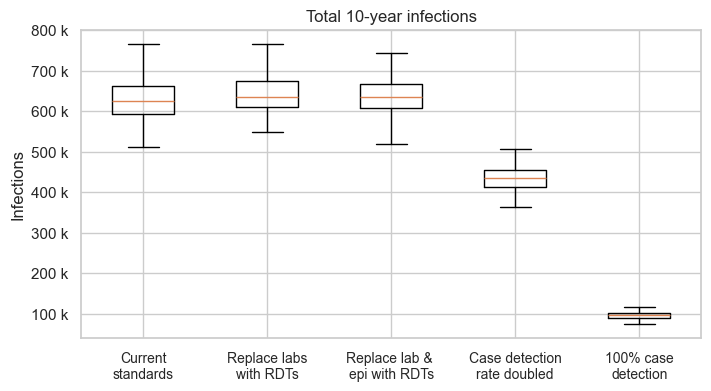

Figure S13: (Top) Distribution of total infections per outbreak (Bottom) distribution of total infections national over 10-years. Replacing laboratory confirmation with RDTs increases median total infections due to the cases missed and later outbreak detection, meaning delayed reactive vaccine campaigns. Increasing RDT coverage decreases total infections due to earlier outbreak detection resulting in faster application of reactive vaccination campaigns.

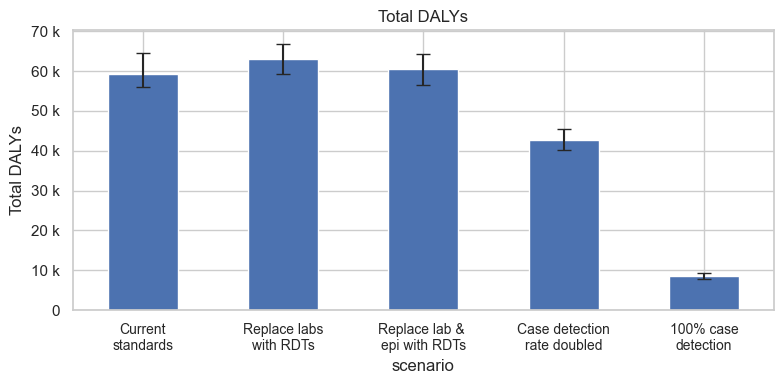

Figure S14: Total DALYs over a 10-year period. Bar height represents the median of all simulations, and the error bars cover the interquartile range.

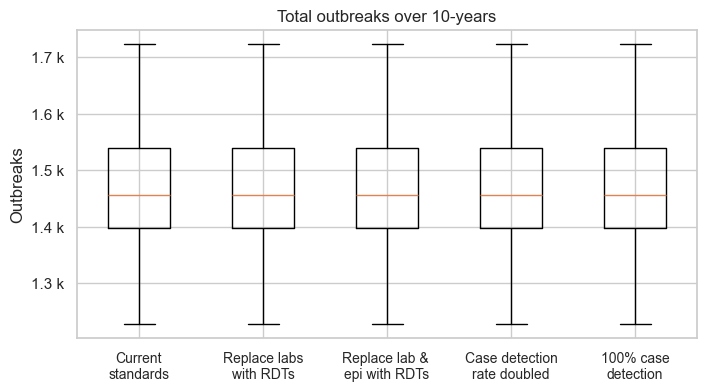

Figure S15: Distribution of total number of outbreaks over 10-years. Outbreaks are sampled randomly from the distribution of monthly outbreak from outbreak dataset, hence scenarios have no impact on the total number of outbreaks over 10-years.

Appendix E Consolidated Health Economic Evaluation Reporting Standards (CHEERS) Checklist

| **Topic** | **No.** | **Item** | **Location where item is reported** |
| --- | --- | --- | --- |
| **Title** |  |  |  |
|  | 1 | Identify the study as an economic evaluation and specify the interventions being compared. | Page 1 |
| **Abstract** |  |  |  |
|  | 2 | Provide a structured summary that highlights context, key methods, results, and alternative analyses. | Page 3 |
| **Introduction** |  |  |  |
| **Background and objectives** | 3 | Give the context for the study, the study question, and its practical relevance for decision making in policy or practice. | Page 4 |
| **Methods** |  |  |  |
| **Health economic analysis plan** | 4 | Indicate whether a health economic analysis plan was developed and where available. | Page 8, Section 2.9 |
| **Study population** | 5 | Describe characteristics of the study population (such as age range, demographics, socioeconomic, or clinical characteristics). | Page 5, Section 2.1 |
| **Setting and location** | 6 | Provide relevant contextual information that may influence findings. | Page 5, Section 2.1 |
| **Comparators** | 7 | Describe the interventions or strategies being compared and why chosen. | Page 7, Section 2.7 |
| **Perspective** | 8 | State the perspective(s) adopted by the study and why chosen. | Page 8, Section 2.8 |
| **Time horizon** | 9 | State the time horizon for the study and why appropriate. | Page 8, Section 2.8 |
| **Discount rate** | 10 | Report the discount rate(s) and reason chosen. | Page 8, Section 2.8 & 2.9 |
| **Selection of outcomes** | 11 | Describe what outcomes were used as the measure(s) of benefit(s) and harm(s). | Page 8, Section 2.9 |
| **Measurement of outcomes** | 12 | Describe how outcomes used to capture benefit(s) and harm(s) were measured. | Page 8, Section 2.9 & 2.10 |
| **Valuation of outcomes** | 13 | Describe the population and methods used to measure and value outcomes. | Pages 5-10 |
| **Measurement and valuation of resources and costs** | 14 | Describe how costs were valued. | Page 8, Section 2.8 |
| **Currency, price date, and conversion** | 15 | Report the dates of the estimated resource quantities and unit costs, plus the currency and year of conversion. | Page 8, Section 2.8 |
| **Rationale and description of model** | 16 | If modelling is used, describe in detail and why used. Report if the model is publicly available and where it can be accessed. | Pages 5-7 |
| **Analytics and assumptions** | 17 | Describe any methods for analysing or statistically transforming data, any extrapolation methods, and approaches for validating any model used. | Pages 5-10 |
| **Characterising heterogeneity** | 18 | Describe any methods used for estimating how the results of the study vary for subgroups. | NA |
| **Characterising distributional effects** | 19 | Describe how impacts are distributed across different individuals or adjustments made to reflect priority populations. | NA |
| **Characterising uncertainty** | 20 | Describe methods to characterise any sources of uncertainty in the analysis. | Page 8, Section 2.10 |
| **Approach to engagement with patients and others affected by the study** | 21 | Describe any approaches to engage patients or service recipients, the general public, communities, or stakeholders (such as clinicians or payers) in the design of the study. | NA |
| **Results** |  |  |  |
| **Study parameters** | 22 | Report all analytic inputs (such as values, ranges, references) including uncertainty or distributional assumptions. | Pages 8-10, Table 1 |
| **Summary of main results** | 23 | Report the mean values for the main categories of costs and outcomes of interest and summarise them in the most appropriate overall measure. | Pages 10-13, Section 3.1 & 3.2 |
| **Effect of uncertainty** | 24 | Describe how uncertainty about analytic judgments, inputs, or projections affect findings. Report the effect of choice of discount rate and time horizon, if applicable. | Pages 10-13, Section 3.1 & 3.2 |
| **Effect of engagement with patients and others affected by the study** | 25 | Report on any difference patient/service recipient, general public, community, or stakeholder involvement made to the approach or findings of the study | NA |
| **Discussion** |  |  |  |
| **Study findings, limitations, generalisability, and current knowledge** | 26 | Report key findings, limitations, ethical or equity considerations not captured, and how these could affect patients, policy, or practice. | Pages 13-14 |
| **Other relevant information** |  |  |  |
| **Source of funding** | 27 | Describe how the study was funded and any role of the funder in the identification, design, conduct, and reporting of the analysis | Page 1 |
| **Conflicts of interest** | 28 | Report authors conflicts of interest according to journal or International Committee of Medical Journal Editors requirements. | Page 1 |

*From:* Husereau D, Drummond M, Augustovski F, et al. Consolidated Health Economic Evaluation Reporting Standards 2022 (CHEERS 2022) Explanation and Elaboration: A Report of the ISPOR CHEERS II Good Practices Task Force. Value Health 2022;25. <doi:10.1016/j.jval.2021.10.008>
